# Blood lipid density decreases at the early-stage of stomach adenocarcinoma

**DOI:** 10.1101/2023.03.01.23286666

**Authors:** Om Prakash, Feroz Khan

**Affiliations:** Technology Dissemination and Computational Biology Division, CSIR-Central Institute of Medicinal & Aromatic Plants, P.O.-CIMAP, Kukrail Picnic Spot Road, Lucknow-226015 (Uttar Pradesh), India

**Keywords:** Cancer, Disease, Early, Prediction, Stomach

## Abstract

**Background:** Induction of cancer creates many molecular to physiological changes in the human body. Observation of these variations between normal & diseased conditions become the basis of disease diagnosis. Present study analyzed blood lipid profile, relative to changes in genotype, at the early stage of stomach adenocarcinoma.

**Materials and Method:** Present study was based on establishment of relationship between genotype to phenotype. Genotypic features were collected through RNAseq analysis, which was further mapped with phenotypic expression in the form of blood lipid profile.

**Results:** To observe the significance difference between phenotypic expressions of normal and cancerous condition, gene signatures from multiple sources of studies were mapped with blood lipid profile including: Total Cholesterol, LDL Cholesterol, HDL Cholesterol, Triglycerides, Non-HDL-C, and TG to HDL ratio. Significance difference found between phenotypic expression of normal and cancerous condition.

**Conclusion:** Through multi-signature-based population observation, it was found that blood-lipid density decreases at the early-stage of stomach adenocarcinoma. Further, blood-lipid profile can be used for early disease prediction of stomach adenocarcinoma as well as other cancer types.

## INTRODUCTION

Stomach adenocarcinoma (STAD) is one of the leading causes of deaths in the world. Lack of early diagnosis is also a major barrier behind this situation. Although, diagnosis through biomarker genes linked with extracellular matrix and platelet-derived growth factor are suggested in early detection of STAD (Tan et.al., 2021). Besides this, CDCA7-regulated inflammatory mechanism (Guo et.al., 2021), angiogenesis-related lncRNAs (Han et.al., 2021), PLXNC1 an immune-related gene (Ni et.al., 2021), and Ferroptosis-related gene etc. are also claimed for early detection of STAD. Different stages of STAD are also available through the prognostic model using the TCGA database. But early diagnosis through pathological blood parameters is not much explored. Various combinations of gene-signatures still remain to explore, which creates an opportunity to reveal aspects for early detection of STAD.

Early prediction of cancer before reaching a life-threatening stage is a challenging problem. Observation of clinicopathological parameters of blood may open a feasible solution to overcome this problem. Blood lipid is a clinicopathological feature which is used to observe disease status. Lipid profile is known to be useful in detection of multiple cancers (Kok et.al, 2011). Lipid analogs are also known as key regulators of tumorigenesis (Dong et.al., 2020). Such studies motivate the observation of cancer in relation to lipid profile (Xiong et.al., 2021). Impact of chemotherapy can also be seen at the blood lipid profile. Hormonal treatment of cancer also shows impact on the blood lipid level (Bundred et.al., 2005). Therefore, blood profile is observed before chemotherapy (Xu et.al., 2020). Researches are being performed on mortalities caused in cancer in relation to blood lipid level and cancer. Such studies consider variation in blood lipid profile (Yang et.al., 2021). Blood lipid level is also observed along with blood glucose and extent of inflammation, as in case of ovarian cancer (Li et.al., 1019). All these studies indicate abnormalities in blood lipid profiles in association with occurrence of cancer (Halton et.al., 1998). Therefore, blood lipid profile is regularly estimated, if a cancer patient is under endocrine therapies (Engan T, 1996). Other studies like phospholipid level & peroxidation estimation have come to the knowledge for malignant neoplasm (Kotrikadze et.al., 1987). Drug response studies on blood profile are also available for tamoxifen, endoxifen and 4-hydroxyTamoxifen (Siqueira et.al., 2021). In totality, potential correlation exists between blood lipid and cancer (Jin et.al., 2020). Diagnostic values are also considerable in combination of tumor markers and blood lipid (Jiang et.al., 2021). These studies provide a ground for establishing a relationship between genotype (tumor markers) and phenotype (blood lipid profile) for observation of import of variation in blood lipid and the cancer status.

RNA expressions are the smallest observation level in several areas of clinical diagnosis (Dube et.al., 2019), including cancer (Lorenzi et.al., 2019). Various categories of RNA are used in cancer diagnosis (Li et.al., 2021; Ruan et.al., 2020). Due to population discrimination capacity of RNA biomarkers, transcriptome sequencing is used in diagnosis of disorders (Akula et.al., 2021). RNA based studies are also known for hepatocellular carcinoma (Chen et.al., 2018), colorectal cancer (Mamelli et.al., 2021), pathway detection (Wu et.al., 2021), prostate tumor (Xia et.al., 2018), and breast cancer (Liu et.al., 2021) etc. More refined, RNA expression profiles are also used for early diagnosis of disease. Early diagnosis is directly linked with development of biomarker signature (i.e., gene set) with population discrimination capacity (Yang et.al., 2020). Such studies provide clinically relevant ground for accessing RNA expressions as classified genotypic features for early detection of cancer.

Genotype-to-phenotype relation is used also for linking lipid metabolism with cancer related genes (Kale et.al., 2020). Patterns in phenotype features can be traced-out by repeated observations in reference to multiple genotypes from different sources. Use of different sources ensures the random sampling of genotypes. Significant observation in pattern modulation in cancer stage, relative to normal, may suggest a feasible pattern for diagnosis at the level of population discrimination. T-test with significant P-value <0.05 is used for identification of null-hypothesis rejected pattern for disease diagnosis. In the present study, variation in blood lipid profile (as phenotype) was linked with gene signature (as genotype) derived for stomach adenocarcinoma, to predict the possible changes/ status of cancer. Inference from this process can be directly used during early diagnosis of stomach adenocarcinoma.

## MATERIALS AND METHOD

Observation of variation in phenotypic expression, at clinicopathological level, for early detection of cancer, needs a genotypic relationship with phenotypic expression. Blood lipid profile was used as a phenotypic expression; while genotypic expression was gene signatures. The method implemented AI-guided relation between blood lipid-profile (clinicopathological phenotype) and expression of gene signature. Workflow behind the protocol is (Figure 1):

**Figure 1.**
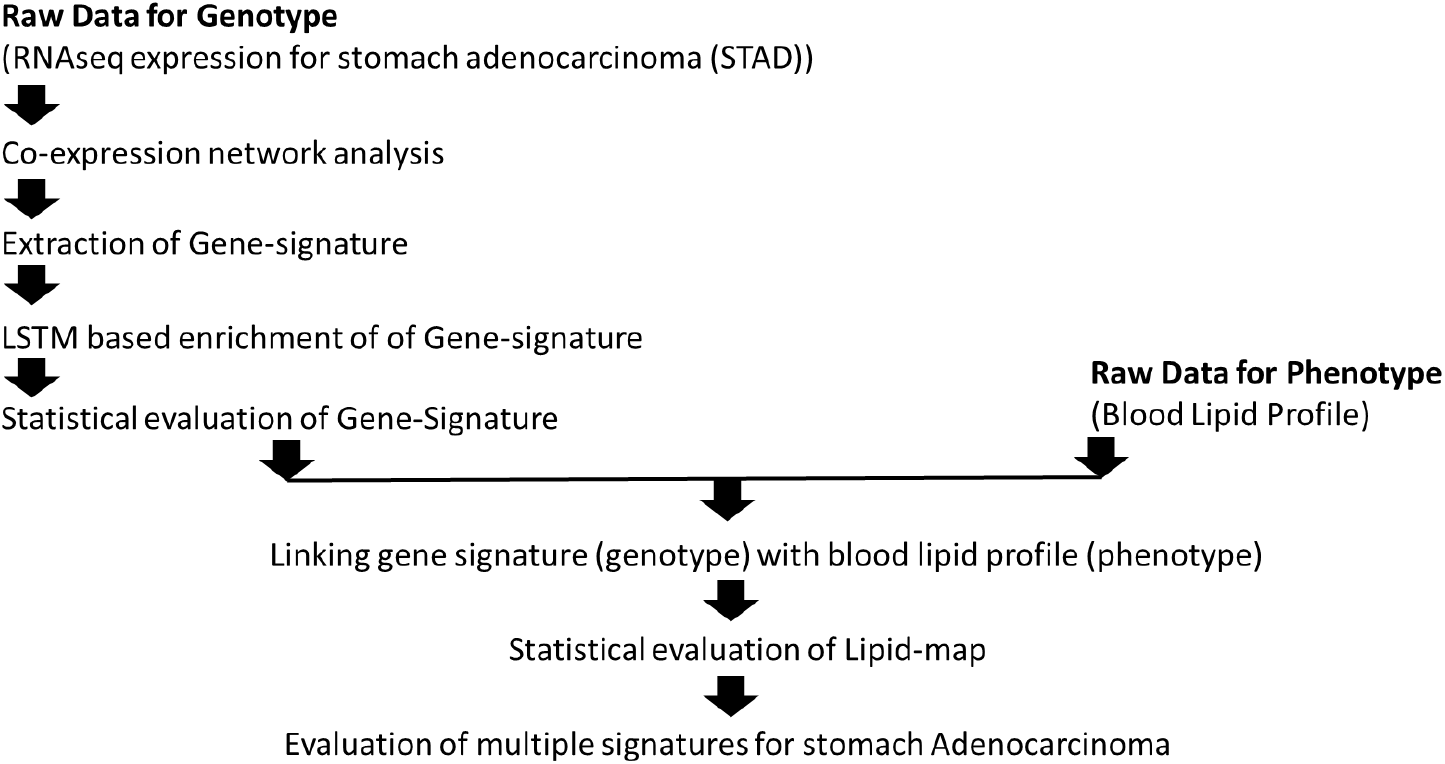
Workflow for assessment of blood lipid density at the early-stage of stomach adenocarcinoma based on blood-lipid profile as clinicopathological feature

### Genotype features

Genotype features were gene-sets with capacity to discriminate diseased population from normal. Classified RNA expressions, for stomach adenocarcinoma, collected as transcriptome sequencing output from the population, 408 independent tumor samples and 211 matching normal tissue samples, were available at TCGA database. Details for plasma containing co-expression of differentially expressed genes (pDEGs) accessed from doi.org/10.1038/s41598-021-87037-w. Further enrichment of co-expression processed through simulation of LSTM systems model. Simulation of system model performed through Long-Short-Term-Memory Recurrent Neural Network (LSTM-RNN) model (implemented in R). Stability of system model accessed through Jacobian matrix. Identified gene-signatures evaluated on the ground of population discrimination capacity through survival plot. LogRank p-value < 0.05 considered as threshold for significant gene-signature.

### Phenotype features

Prior studies indicated lipid-profile for observation during cancer. Such observations may support in overcoming hurdles before early detection of cancer seeded at molecular level. Clinicopathological parameters used as phenotypic features: Total Cholestrol: 200-239(mg/dL); LDL Cholestrol: 130-159 (mg/dL); HDL Cholestrol: 40-60 (mg/dL); Triglycerides: 150-199 (mg/dL); Non-HDL-C: 130-159 (mg/dL); and TG to HDL ratio: 3.0-3.8 (mg/dL).

### AI-guided map for genotype-vs-phenotype

LSTM enriched gene signature (genotype) mapped with lipid-profile (phenotype) of blood. Total 740 data-points used to map lipid-profile with gene-signature.

Since RNA gene expressions were classified, therefore classified gene expression for gene-signature was further used for generating classified mapped lipid-profile dataset. The classified genotypic & phenotypic dataset sets were further used for development of map-model for transformation of blood profile into gene expression. The map-model was developed on the framework of feed-forward backpropagation Artificial Neural Network (FFBP-ANN). Inputs of 06 nodes from lipid-profiles were mapped with outputs of 5 nodes from enriched pDEGs. Therefore, population discrimination capacity was also evaluated for mapped lipid profile. Total 740 (370 (normal) + 370 (cancer)) data-points were used to map lipid-profile with gene-signature. The mapped lipid-data points were evaluated for its discrimination capacity through two class multiple perceptron classification model evaluation. Statistical analysis was performed to evaluate the classification model through ROC plot and AUC calculation.

### Assessment of phenotype (blood-lipid-profile) in reference to multiple gene signatures

To find out the possible variation in phenotype (blood-lipid profile) during the early stage of Adenocarcinoma, blood lipid profile was evaluated in reference to multiple gene signatures. The identified inference can be used as a rule.

## RESULTS AND DISCUSSION

### Gene-signature (genotype features)

Processing of classified RNA expression data, resulted in 62 differentially expressed genes. Out of which, 39 DEGs were found in plasma proteomes. Gene co-expression network analysis resulted in signatures with capacity of discrimination between populations of diseased & non-diseased samples. Only 11 genes (AFF3, APBB1, C5, CHRD, COL4A5, EEF1A2, ZBTB16, IL1RL1, GFRA3, ZNF662, and MAGED4B), out of 39, found to be involved in defining signature. Signature showed population discrimination capacity with p-value of 6E-05. Furthermore, 11 genes were used for development of LSTM-guided systems models. After simulation, out of 11 genes, only 05 genes (AFF3, APBB1, C5, CHRD, and COL4A5) were found to be involved in successful simulation. The enriched five genes also showed population discrimination with a p-value of 0.0046. It is shown that the most significant impact of gene signature on stomach adenocarcinoma is due to genes involved in extracellular matrix organization. Prior study also showed compliance with present results (Tan et.al., 2021). These 05 genes were further used for establishing relation with blood profile (https://doi.org/10.1101/2023.02.14.23285899).

### Linking gene-signature with blood lipid profile

Six blood profile parameters (Total Cholesterol: 200-239(mg/dL); LDL Cholesterol: 130-159 (mg/dL); HDL Cholesterol: 40-60 (mg/dL); Triglycerides: 150-199 (mg/dL); Non-HDL-C: 130-159 (mg/dL); and TG to HDL ratio: 3.0-3.8 (mg/dL)) used for phenotypic observation. Expression of gene signature further processed for mapping with profile of blood. The genotype-phenotype map used for development of ANN classification model, including 05 gene-expression and blood lipid-profile. Normal-to-Cancer discrimination capacity of lipid profile evaluated through multi-perceptron ANN classification model. To evaluate performance of mapping lipid-profile, multiple perceptron model architecture of 6-4-2 implemented with learning rate 0.3 and momentum of 0.2 for 500 epochs. Model established with AUC of 0.999 (Supplementary Table 1, 2 & 3). Ten-fold cross-validation of the model showed accuracy of 99.46%.

### Estimation of Possible variation in blood-lipid profile during early stage of Adenocarcinoma

Since in the case of early detection of cancer; a normal healthy person will be assumed a subject of observation. In that condition, Blood profile will also lie in pathological normal states. So now in that case, how can we detect the possible future coming disease? Can the primary-waves of the disease be detectable? Here, we have to assume that disease-mechanism has already started in the physiology of the body, but phenotypic expressions will come in the near future, which is absent in the present time. Since, genotypic features, for stomach adenocarcinoma, are identified; therefore, genotypic fluctuations can be seen on the frame of pathological parameters, for example Lipid-profile. To know the pattern in phenotypic fluctuation, blood lipid profile was evaluated in reference to multiple gene signatures from prior studies too (Table 1, 2). Phenotypic observations were analyzed through t-test. It was found that there was a significant fall in density of lipid in blood in cancerous conditions than normal. Other prior studies, on lipids in relation to progression of cancer, also showed that there is relation between the variations in level of cholesterol during progression of cancer (Mayengbam, Shyamananda Singh et al. 2021). The possible reason for the fall in cholesterol is that cancer cells consume cholesterol for rapid cell division. Since rapid cell division capacity develops at the early stage of cancer, therefore variation in level of lipid/cholesterol may be an observatory point for early detection of cancer. Since blood sampling is a key part of clinicopathological observation, and it is also linked with supply chain of nutrition for cell division; therefore blood-lipid profile may be a key component for early detection of cancer. Therefore, this result can be considered as a general rule for early detection of cancer (Table 1, 2 & 3).

**Table 1.** Signatures considered into study

| **Signature Sl. No.** | **Gene Signature** | **Reference** |
| --- | --- | --- |
| **Sig-0** | "AFF3", "APBB1", "C5", "CHRD", "COL4A5", "TOP2B", "SMC4" | In-house identified signature |
| **Sig-1** | "GSTA2", "POLD3", "GLA", "GGT5", "DCK", "CKMT2", "ASAH1", "OPLAH", "ME1", "ACYP1", "NNMT", "POLR1A", "RDH12" | Front. Oncol., 21 June 2021 Sec. Gastrointestinal Cancers<br>https://doi.org/10.3389/fonc.2021.612952 |
| **Sig-2** | "ACTA2", "AREG", "ASCC2", "BEST1", "BRCA1", "CREBBP", "DDX5", "EP300", "ESR1", "FAM96A", "FHL2", "GNL3", "HIPK2", "HSF1", "IGSF9", "JUN", "MSH6", "NCOA6", "NCOA6IP", "PARP1", "PAWR", "PCNA", "PML", "PPP2R5A", "RPA1", "SMAD3", "SMARCA4", "TP53", "TP63", "WRN", "WT1", "WTAP" | Cheong, JH., Wang, S.C., Park, S. et al. Development and validation of a prognostic and predictive 32-gene signature for gastric cancer. Nat Commun 13, 774 (2022).<br>https://doi.org/10.1038/s41467-022-28437-y |
| **Sig-3** | "PURB", "SMC1L1", "DENND1B", "PDCD4" | https://doi.org/10.3892/ol.2018.8577 |
| **Sig-4** | "ABCE1", "ADH1C", "ADNP", "ALDH6A1", "APOC1", "APOE", "ATP13A3", "BAZ1A", "BCAR3", "CAPRIN1", "CBFB", "CCT2", "CCT6A", "CEP55", "CHORDC1", "COL6A3", "CPXM1", "CXCL1", "CXCL10", "ECHDC2", "ENC" | https://doi.org/10.1016/j.ebiom.2020.103023 |
|  | 1","EPHB4","ETFDH","FGFR4","FHOD1","GABBR1","GART","INHBA","KAT2A","KLF4","LAMC2","LIMK1","LRRC41","MCM2","MMP14","NCL","NOL8","OSMR","P4HA1","PDP1","PGRMC2","PNO1","PRC1","PRR7","PTGS1","SCNN1B","SLC12A9","SLC20A1","SMS","TCERG1","TGS1","TNFAIP2","TUBB" |  |
| **Sig-5** | "CD209", "TOP3B", "BCL2", "MCM2", "RAD9A", "IL10", "HLA-DMB", "MCM10", "PTPRC", "FANCM", "ADORA2A", "MS4A1", "FANCG", "CD3G", "DSCR6", "TBX21", "ZWILCH", "IL17A", "FCGR3A" | https://gut.bmj.com/content/71/4/676 |
| **Sig-6** | "VCAN","EFNA3","ADH4","CLDN9" | Ann Transl Med 2022;10(18):1010 \| https://dx.doi.org/10.21037/at |

**Table 2.** Phenotypic expressions in blood lipid-profile in relation of gene-signatures for Gastric cancer considered during study.

| Signature<br>Sl. No. | Phenotypic expressions in blood lipid-profile |  |  |  |  |  |  |
| --- | --- | --- | --- | --- | --- | --- | --- |
| Sig-0 |  |  |  |  |  |  |  |
|  | Normal: | 214.2378 | 174.1326 | 47.65243 | 270.8775 | 169.9518 | 3.69331 |
|  | Tumor: | 207.9712 | 170.757 | 45.91087 | 227.1245 | 149.5762 | 3.34872 |
| Sig-1 |  |  |  |  |  |  |  |
|  | Normal<br>: | 235.3203 | 146.494 | 49.63761 | 186.109 | 147.512 | 4.04113 |
|  | Tumor: | 221.4865 | 143.7998 | 46.73759 | 187.4013 | 150.1777 | 3.914579 |
| Sig-2 | signature could not run |  |  |  |  |  |  |
| Sig-3 | signature could not run |  |  |  |  |  |  |
| Sig-4 |  |  |  |  |  |  |  |
|  | Normal: | 346.6813 | 169.8594 | 58.09198 | 251.5707 | 241.0234 | 6.303563 |
|  | Tumor: | 279.8497 | 157.0263 | 50.49587 | 202.5042 | 196.4386 | 4.701655 |
| Sig-5 | signature could not run |  |  |  |  |  |  |
| Sig-6 |  |  |  |  |  |  |  |
|  | Normal: | 219.0083 | 142.7449 | 57.39752 | 170.4471 | 156.4613 | 3.182708 |
|  | Tumor: | 215.6043 | 144.6829 | 45.25915 | 172.4644 | 144.9133 | 3.213511 |

**Table 3.** T-Test: Paired Two Sample for Means for Normal and Tumor lipid profile from different gene signatures.

|  | *Normal* | *Tumor* |
| --- | --- | --- |
| Mean | 144.6847 | 132.3067 |
| Variance | 9393.133 | 7810.059 |
| Observations | 6 | 6 |
| Pearson Correlation | 0.998567 |  |
| Hypothesized Mean Difference | 0 |  |
| df | 5 |  |
| t Stat | 3.070027 |  |
| P(T<=t) one-tail | 0.013891 |  |
| t Critical one-tail | 2.015048 |  |
| P(T<=t) two-tail | 0.027783 |  |
| t Critical two-tail | 2.570582 |  |

## Conclusions

There are many computational approaches to understand the relationship between blood lipid and cancer. The relation between gene signature from population and blood lipid profile has never been tried for early detection of cancer. Therefore, in this article, we performed a computational study for understanding this relation at the early stage of cancer. In this regard, RNA-seq expression, from normal and diseased populations, is processed for stomach adenocarcinoma. We collected phenotype and genotype representation for the early stage of stomach adenocarcinoma, and performed gene-signature identification through co-expression and DEGs analysis. Gene-signature’s capacity evaluated for discrimination between healthy and diseased population. Gene-signature expression mapped with blood lipid profile. Finally, variation in blood-lipid profile, during the early stage of adenocarcinoma, was observed for multiple gene-signatures reported in prior studies. Significant fall in density of blood-lipid profile found during the early stage of stomach adenocarcinoma. This inference from the study can be very useful for early detection of stomach adenocarcinoma. Furthermore, more studies can be performed for other cancer types also.

## Data Availability

All data produced in the present work are contained in the manuscript

## Acknowledgement

Authors are thankful to the Director, CSIR-Central Institute of Medicinal & Aromatic Plants (CIMAP), Lucknow, India for infrastructure & research facilities support. Author OP is thankful to the Indian Council of Medical Research (ICMR), New Delhi, India for financial support through RA fellowship (Award letter no. BMI/11(12)/2020, dated: 04/02/2021). Author OP is also thankful to Prof. Thiol Gross, Helmholtz Institute for Functional Marine Biodiversity (HIFMB) at University of OLDENBURG, Germany and Dr. Amit Singh, Chennai Mathematical Institute (CMI) Chennai, India for fruitful suggestions on evaluation of stability of time series systems model. The CSIR-CIMAP publication number of this manuscript is CIMAP/PUB/ 2023/24.

## Conflict of interest

There is no conflict of interest.

---

**Supplementary Table 1.** Statistics showing evaluation through 10-fold cross validation of two-class classification model for evaluation of discrimination capacity of lipid-profile

|  | TP | FP | Precision | Recall | F-Measure | MCC | ROC area | PRC area | Class |
| --- | --- | --- | --- | --- | --- | --- | --- | --- | --- |
|  | 0.997 | 0.008 | 0.992 | 0.997 | 0.995 | 0.989 | 1 | 1 | T |
|  | 0.992 | 0.003 | 0.997 | 0.992 | 0.995 | 0.989 | 1 | 1 | N |
| Weighted Avg. | 0.995 | 0.005 | 0.995 | 0.995 | 0.995 | 0.989 | 1 | 1 |  |
| Correctly Classified Instances 736 99.4595 % |  |  |  |  |  |  |  |  |  |
| Incorrectly Classified Instances 4 0.5405 % |  |  |  |  |  |  |  |  |  |
| Kappa statistic 0.9892 |  |  |  |  |  |  |  |  |  |
| Mean absolute error 0.0091 |  |  |  |  |  |  |  |  |  |
| Root mean squared error 0.0631 |  |  |  |  |  |  |  |  |  |
| Relative absolute error 1.8151 % |  |  |  |  |  |  |  |  |  |
| Root relative squared error 12.6272 % |  |  |  |  |  |  |  |  |  |
| Total Number of Instances 740 |  |  |  |  |  |  |  |  |  |

**Supplementary Table 2.** Phenotypic expression related with Normal cell

| Sig1 | Sig2 | Sig3 | Sig4 | Avg (Normal Signature) |
| --- | --- | --- | --- | --- |
| 214.2378 | 235.3203 | 346.6813 | 219.0083 | 253.8119 |
| 174.1326 | 146.494 | 169.8594 | 142.7449 | 158.3077 |
| 47.65243 | 49.63761 | 58.09198 | 57.39752 | 53.19489 |
| 270.8775 | 186.109 | 251.5707 | 170.4471 | 219.7511 |
| 169.9518 | 147.512 | 241.0234 | 156.4613 | 178.7371 |
| 3.69331 | 4.04113 | 6.303563 | 3.182708 | 4.305178 |

**Supplementary Table 3.** Phenotypic expression related with Tumor cell

| Sig1 | Sig2 | Sig3 | Sig4 | Avg (Tumor Signature) |
| --- | --- | --- | --- | --- |
| 207.9712 | 221.4865 | 279.8497 | 215.6043 | 231.2279 |
| 170.757 | 143.7998 | 157.0263 | 144.6829 | 154.0665 |
| 45.91087 | 46.73759 | 50.49587 | 45.25915 | 47.10087 |
| 227.1245 | 187.4013 | 202.5042 | 172.4644 | 197.3736 |
| 149.5762 | 150.1777 | 196.4386 | 144.9133 | 160.2765 |
| 3.34872 | 3.914579 | 4.701655 | 3.213511 | 3.794616 |

## Notes

### Competing Interest Statement

The authors have declared no competing interest.

### Funding Statement

Author Om Prakash is thankful to the Indian Council of Medical Research (ICMR), New Delhi, India for financial support through a Research Associate fellowship (Award letter no. BMI/11(12)/2020, dated: 04/02/2021).
The authors utilized research infrastructure at the Institute of Medicinal & Aromatic Plants (CIMAP), Lucknow, India.

### Author Declarations

Data sources (publically available) considered during the study: (1) The Cancer Genome Atlas, The GDC data portal https://portal.gdc.cancer.gov/. (2) Gene Expression Profiling Interactive Analysis database (http://gepia2.cancer-pku.cn/#index). (3) cBioPortal for Cancer Genomics (https://www.cbioportal.org/). (4) Pubmed (https://pubmed.ncbi.nlm.nih.gov/).

